# Protocol for a cross-sectional study: Effects of a Multiple Sclerosis Relapse Therapy with Methylprednisolone on Offspring Neurocognitive Development and Behaviour (MS-Children)

**DOI:** 10.1101/2021.11.15.21266211

**Authors:** Valeska Kozik, Matthias Schwab, Sandra Thiel, Kerstin Hellwig, Florian Rakers, Michelle Dreiling

**Author notes:** **<u>Corresponding author:</u>** Valeska Kozik, Department of Neurology, Jena University Hospital, Am Klinikum 1, 07747 Jena, Germany. authors contributed equally (Florian Rakers, Michelle Dreiling).

## Abstract

**Introduction:** Multiple Sclerosis (MS) is the most common neuroimmunological disease in women of childbearing age. Current MS therapy consists of immunomodulatory relapse prevention with disease-modifying therapies (DMTs) and acute relapse therapy with the synthetic glucocorticoid (GC) methylprednisolone (MP). As most DMTs are not approved for use during pregnancy, treatment is usually discontinued, increasing the risk for relapses. While MP therapy during pregnancy is considered relatively save for the foetus, it may be detrimental for later cognitive and neuropsychiatric function. The underlying mechanism is thought to be an epigenetically mediated desensitisation of GC receptors, the subsequent increase in stress sensitivity, and a GC-mediated impairment of brain development. The aim of this study is to investigate the associations of foetal MP exposure in the context of MS relapse therapy with later cognitive function, brain development, stress sensitivity, and behaviour.

**Methods and analysis:** 80 children aged 8 to 18 years of mothers with MS will be recruited. 40 children, exposed to GC in utero will be compared to 40 children without foetal GC exposure. The intelligence quotient will serve as primary outcome. Secondary outcomes will include attention, motor development, emotional excitability, Attention-Deficit Hyperactivity Disorder-related symptoms, and behavioural difficulties. The Trier Social Stress Test will test stress sensitivity, EEG and MRI will assess functional and structural brain development. To determine underlying mechanisms, DNA methylation of the GC receptor gene and the H19/IGF2 locus and changes in the microbiome and the metabolome will be investigated. Primary and secondary outcomes will be analysed using linear regression models. Time-variant outcomes of the stress test will be analysed in two mixed linear models exploring overall activity and change from baseline.

**Ethics and dissemination:** This study was approved by the participating institutions’ ethics committees and results will be presented in accordance with the STROBE 2007 Statement.

**Trial registration:** https://clinicaltrials.gov/ct2/show/NCT04832269?id=ZKSJ0130

## INTRODUCTION

### Background

Multiple sclerosis (MS) is one of the most common neurological diseases in young people, affecting approximately 2.3 million patients worldwide [1]. Approximately two thirds of these patients are women, mainly of childbearing age [2]. Therapy for MS consists of disease-modifying therapy (DMT) for relapse prophylaxis with immunomodulatory substances and relapse therapy with methylprednisolone (MP). As most immunomodulators are not approved for use during pregnancy, treatment is usually discontinued with a subsequent risk increase for relapses [3]. Although the hormonal changes during pregnancy decrease the risk of relapses significantly during the third trimester, risk for relapses especially early during the pregnancy is comparable to the prepartum period and thus not an uncommon event [4].

European and US guidelines recommend the use of high-dose MP to treat severe, disabling relapses anytime during pregnancy with a risk-benefit-evaluation between gestational weeks 8 and 11 [5]. This treatment is considered relatively harmless for the foetus since no severe physical development disorders have been found at birth [3, 6]. However, studies with offspring antenatally exposed to the synthetic GC betamethasone (BM) to enhance foetal lung maturation due to a threat of premature labour suggest a risk for increased stress sensitivity and neuropsychiatric disorders in later life [7, 8]. Remarkably, the treatment regime used in obstetric indications consists of only 2 × 8-12 mg BM 24 h apart as compared to the 500-1000 mg MP over 3 – 5 days used to treat an MS relapse. Considering the approximately six-fold higher biological activity of BM compared with MP [9], the total dose of GC in the treatment of MS relapses is up to 50-fold higher than for obstetric indications. In a previous study from our workgroup, which is the basis for the present study, we could show that antenatal exposure to 2 × 8 mg BM to enhance foetal lung maturation resulted in multidimensional changes in neurodevelopment and stress-sensitivity in full-term offspring at 8 – 9 years of age compared to children not exposed to BM [10, 11]. This included an intelligence quotient (IQ), which was on average 10 points lower in the exposed group than in the non-exposed group, changes of autonomic activity, attention-deficit hyperactivity disorder (ADHD)-like symptoms and changes in electrocortical activation during a cognitive task. In a follow-up study with 16-year-olds antenatally exposed to BM, we found changes in measures of structural brain development, specifically alterations in gyrification and a reduced cortical thickness in MRI scans (publication in preparation). Combined, these findings have led to a change in German obstetric guidelines already, which now call for a more critical indication for the induction of lung maturation [12, 13].

The adverse long-term effects of antenatal GC exposure may be the result of both a programming of the function of the hypothalamic-pituitary-adrenal axis (HPAA) and maturational effects of GCs on functional and structural brain development. The relationship between adverse environmental influences such as an intrauterine exposure to supraphysiological levels of GCs during critical periods of development and the health of the offspring in later life is the basis of the ‘Foetal Programming’ or ‘Developmental Origins of Health and Disease (DOHaD) hypothesis’ [14, 15]. Until the 3^rd^ trimester, the foetus is incapable of synthesising cortisol and relatively protected from maternal cortisol because the placental enzyme 11ß-HSD 2 inactivates 80 – 90 % of maternal cortisol to cortisone [16]. In contrast to cortisol, most synthetic GCs with the exception of MP are no substrate for 11ß-HSD2 and cross the placenta without inactivation [17]. However, even though MP is also a substrate for 11ß-HSD 2, high bolus doses of MP as used in MS-relapse therapy rapidly saturate this enzyme and reach the fetus in pharmacological concentrations [6, 18]. Once in the fetal circulation, even short-term elevated levels of GCs can permanently change patterns of gene expression and cellular function via epigenetic mechanisms [19]. Permanent desensitisation of GC receptors involved in negative feedback regulation of the HPAA via DNA methylation of the promoter region of the GC receptor gene *NR3C1* may lead to a life-long hyperactivity of the HPAA and, thus, to increased stress sensitivity in later life [20, 21]. Hyperactivity of the HPAA is associated with stress-related neuropsychiatric disorders, including anxiety, depression, and ADHD [20, 21].

Increased levels of GCs induce accelerated tissue differentiation and maturation at the expense of tissue growth [22]. While this effect is utilised to induce lung maturation in foetuses at risk of premature birth [23], it occurs in all organ systems and is reflected in a reduced birth weight following antenatal GC treatment [24]. Out of all organ systems, the developing brain is particularly vulnerable to elevated GC concentrations due to its complex sequence of precisely coordinated steps of maturation and its intrinsic plasticity [25], a basic property of the brain enabling learning and adaptation. Supraphysiological concentrations of GCs change foetal brain development via antiproliferative effects on neural stem and progenitor cells [26]. The subsequent changes in neuronal connections and myelination [27, 28] may contribute to the sustained changes in cognitive and cerebral functioning. Increased concentrations of GCs may also affect fetal brain development indirectly *via* changes in the composition of the maternal gut microbiota [29, 30], which in turn modulates the foetal gut microbiota [31, 32]. The gut microbiome communicates with the central nervous system *via* an integrated, bidirectional communication pathway that, amongst others involves inflammatory cytokines, neuromodulators and neurotransmitters [16]. Consequently, disturbances in this communication system, the so called ‘gut-brain’ axis, have been associated with a number of neurological and psychiatric diseases, such as depression, anxiety disorders, and attention deficit hyperactivity disorder [16, 31, 33, 34].

While no studies examining the long-term effects on the offspring exist, current MS guidelines consider relapse therapy using MP during pregnancy safe for the foetus. Based on findings regarding long-term consequences of antenatal BM exposure, we hypothesise however, that this may not be the case. Based on the up to 50-fold cumulative dose of GCs in MS relapse treatment and its more prolonged administration compared to obstetric indications, we aim to investigate whether relapse treatment with MP in MS patients during pregnancy affects structural and functional brain development, cognition and behaviour as well as stress sensitivity in the antenatally exposed offspring.

### Research aims and hypotheses

This study is based on two hypotheses: Foetal exposure to MP during MS relapse therapy leads to epigenetically mediated

a. disturbances in functional and structural brain development and
b. a long-term change in stress sensitivity of the offspring.

The resulting changes in brain function are associated with neuropsychiatric abnormalities as well as cognitive and behavioural changes in later life.

The primary objective of this cross-sectional study is to elucidate associations of antenatal MP exposure with cognitive performance in children aged 8 – 18 years compared to non-exposed children of mothers with MS. As secondary objectives, this study will investigate the association of cognitive development and behavioural problems with underlying changes in stress sensitivity and functional and structural brain development. To clarify potential epigenetic mechanisms underpinning these changes in stress sensitivity and functional and structural brain development, DNA methylation at the promotor region of the GC receptor gene *NR3C1* and at the H19/IGF2 locus, which controls expression of the insulin-like growth factor 2 (IGF2), the major growth hormone during development [35], will be investigated. To elucidate further potential mechanisms, changes in the composition of the microbiome will be examined.

This study is expected to advance the understanding of developmental programming effects of antenatal high-dosage GC exposure. Another major aim of this study is to add data to the debate around evidence-based decision-making regarding (1) the continuation of a DMT during pregnancy *vs*. the potential long-term risks of relapse therapy with MP for the child’s cognitive and neuropsychiatric health in later life, and (2) the risks of relapse treatment with MP for the child *vs*. the risks of an untreated relapse for the mother. There is an accumulating amount of *real world* data on the safety of continuing a DMT during pregnancy primarily with interferons or glatiramer acetate, which may be safer for mother and child than a potentially necessary relapse treatment with MP.

## METHODS AND ANALYSIS

### Study design

This two-centre, observational, cross-sectional study aims to recruit N = 80 children of mothers with MS. The exposed group (n = 40) will consist of children who were antenatally exposed to MP as part of maternal MS relapse therapy, while the control group (n = 40), will consist of children, who were not exposed to MP. Assessment will take place at the University Hospital in Jena (UKJ) and the Ruhr University Bochum (RUB). The first participant was included on 19/10/2020 and the study is anticipated to continue until the end of 2024.

### Sample size

The aforementioned study by some of the authors [10] investigated cognitive performance, as measured by an IQ test, in two groups of 39 healthy children at the age of 8 years. Considering the lower biological activity of MP compared to BM, the cohort in the present study was exposed to a GC-dose up to 50-fold higher than in the study on the effects of BM. The higher dosage of MP will most likely expose a significant effect in two groups of the same size as the previous study.

As the present study uses a different IQ test, the planned sample size is further based on the standardised effect size Cohen’s *d*, as a multiple of the standard deviation. The previous study found a mean difference in IQ score of 10.5 points (SD = 14.0). A two-sample t-test (α = .05) would have sufficient power of 82% (*d* = .71) to 87% (*d* = .75) with two groups of 35 children. This takes into account potential non-evaluability of five participants per group, e.g., in the case of artefacts or attrition. The planned regression models include seven variables, which, based on the *rule of ten* leads to a sufficient number of cases to produce robust results [36].

### Sample characteristics and recruitment

Children aged 8 to 18 years, of any sex and ethnicity are eligible to participate. A physician presents verbal and written information about the study, its aims, the procedure, and data management to the participants and their guardian(s) and answers any remaining questions. Children are included if they speak German and they and their guardian(s) are willing and able to consent to their participation.

Exclusion criteria for both groups comprise preterm birth (before 34^th^ week of gestation), a birth weight below the 5^th^ percentile, and perinatal complications, such as cerebral haemorrhage, neonatal intensive care requiring ventilation, or any further antenatal therapy with GCs beyond MS relapse therapy. Participants are further excluded from the study in case of maternal substance abuse during pregnancy or serious illness in the child which makes assessment impossible, such as an intellectual disability, or long-term medication with GCs, e.g., in asthma.

Recruitment takes place in the UKJ’s close cooperation with the RUB, the host of the *German MS and Pregnancy Registry*, and several other MS centres across Germany. A search algorithm was developed to identify eligible participants for the MP-exposed and the control group within the respective hospital-specific data management systems. Further recruitment efforts are directed towards online advertisements on MS-specific websites and UKJ’s public outreach channels.

### Outcome measures

See tables 1 and 2 for detailed description of outcome measures and scheduling of data collection. The primary outcome measure of this study is the child’s global cognitive ability, as measured by a standardised IQ-test, i.e., the Reynolds Intellectual Assessment Scales and Screening (RIAS) [37]. Secondary behavioural outcomes, such as motor development, attention, emotional excitability, ADHD-associated symptoms, and behavioural difficulties in multiple categories are measured via neuropsychological assessment, and self-, and informant-reported questionnaires. Additional secondary outcomes are obtained during the Trier Social Stress Test for children (TSST-C), an established standardised protocol to test stress sensitivity [38]. These outcome measures comprise of the neuroendocrine (salivary cortisol) and autonomic stress response (salivary alpha-amylase concentration as a measure of sympathetic activity [39]) and heart rate variability (HRV) indices. To capture subjective stress levels during the TSST-C, the participant fills out an anxiety questionnaire [40] before and after the test. As measures of neurodevelopment, activation in the EEG during rest and a cognitive task included in the TSST-C and MRI-based markers of structural brain development are used. With the use of peripheral blood (leukocytes), and buccal mucosa cells, potential epigenetic mechanisms underlying changes in cognition and behaviour as well as in stress sensitivity will be determined (see table 2). For exploratory mechanistic analysis, the composition of the microbiome (stool sample), metabolome (EDTA plasma) and elementome (hair sample) will be examined.

**Table 1:** Basic clinical and demographic data collected.

| Variables | Instruments/Scale | Collection |
| --- | --- | --- |
| Demographic data of participant and guardian(s) | Demographic and socioeconomic data are collected from the participants and their guardian(s) with questionnaires and an interview, including age, sex, height, weight, guardians' education (highest degree), marital status, and chronic health conditions. | NPT day |
| Basic birth data | Birth data for the child (gestational age at birth, | NPT day |
|  | birth weight and length, head circumference at birth) and mother (diabetes mellitus and pre-eclampsia) are collected via an interview and the consultation of medical files. |  |
| MS-specific medical data | To describe the mother's MS status during pregnancy, medical files and the mother are consulted regarding disease course and duration, therapies, and disease progression. | NPT day |
| Stressors (pregnancy) | Based on similar validated instruments, a questionnaire adapted to this study's aims is used to assess stress during the pregnancy. For each trimester, the mother indicates any stressors related to personal, familial, or work life, or noxious substances intake. | NPT day |
| Stressors (child) | A German self-report questionnaire is used to determine stressful life events in the past 12 months of the participant's life (Zürcher Life Events List) [41], involving home life, school, friendships, etc. In the affirmative case, the participant rates the level of stress the event caused on a 5-point scale. | NPT day |

*Note: NPT = Neuropsychological Testing*

**Table 2:**
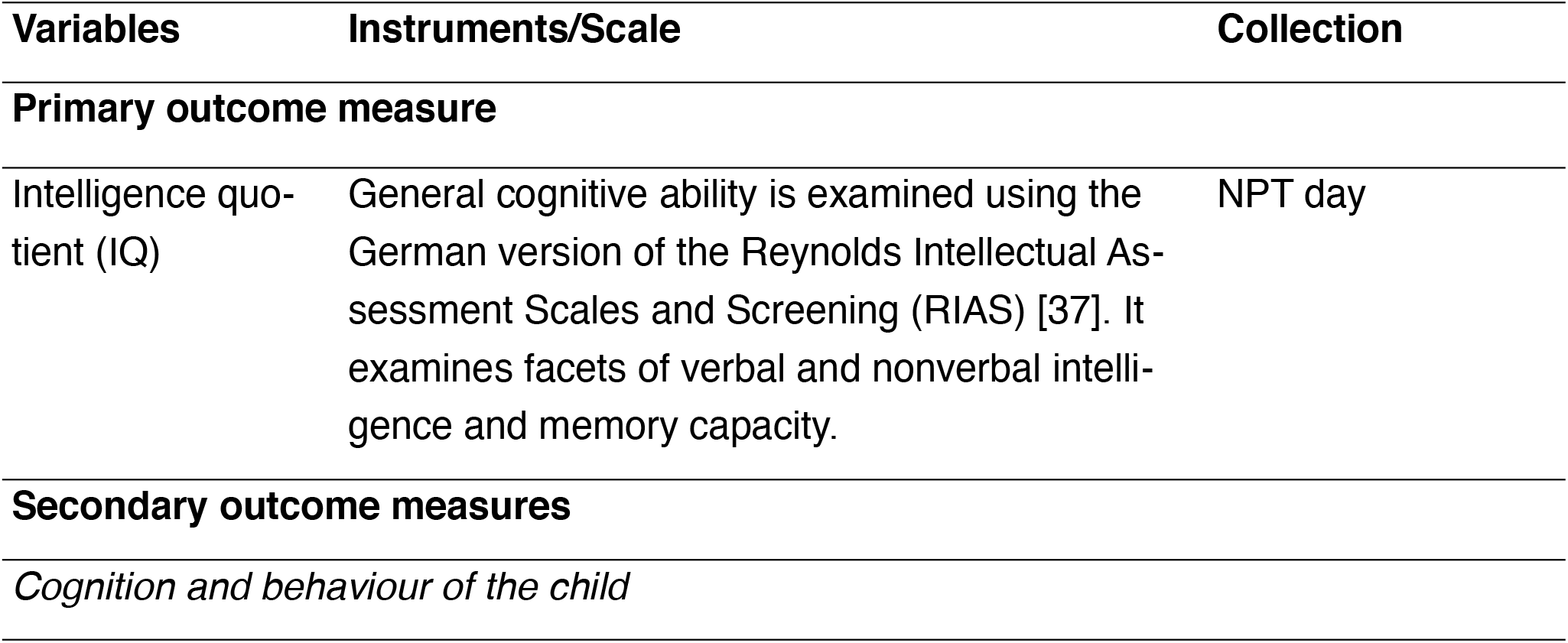
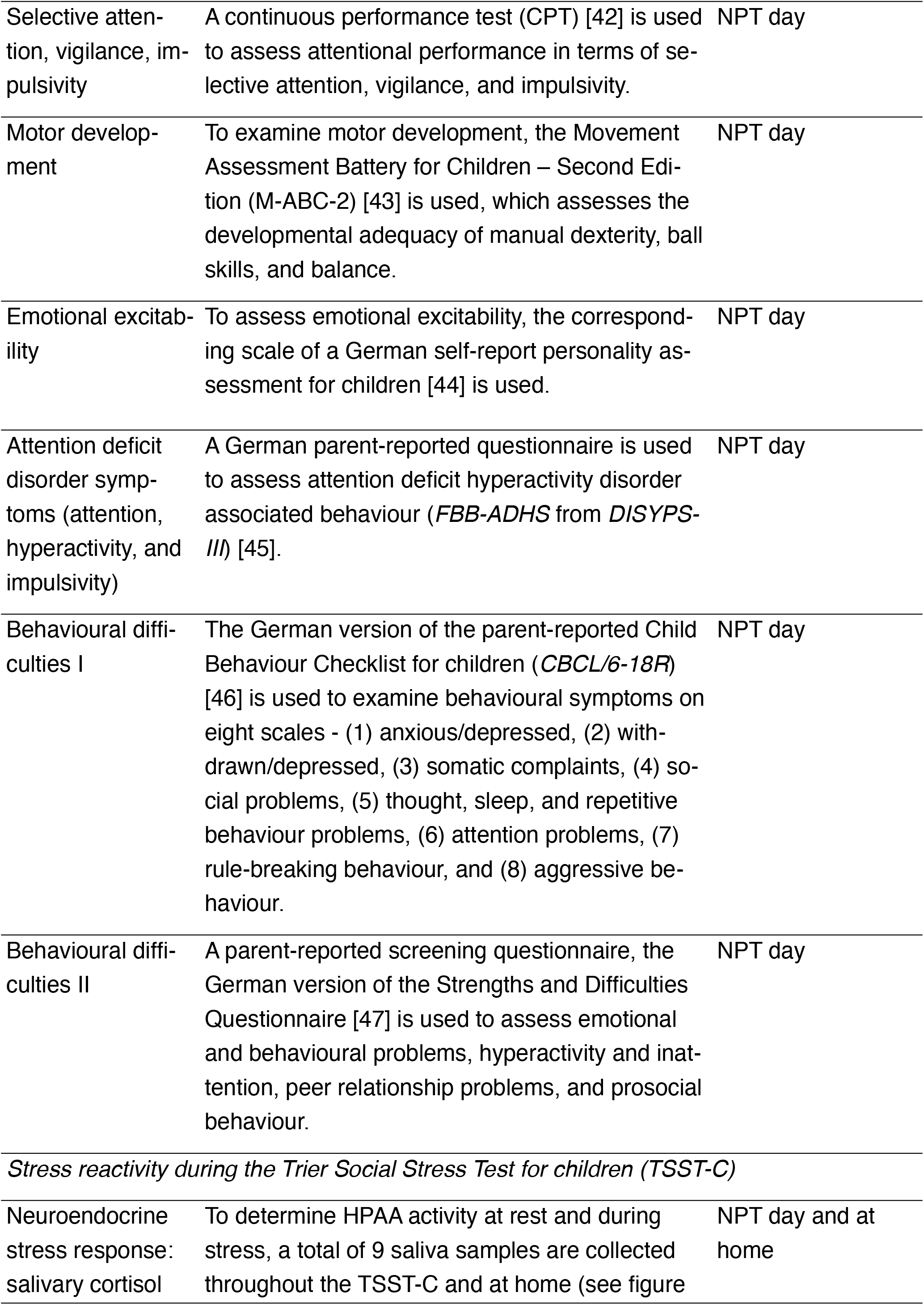
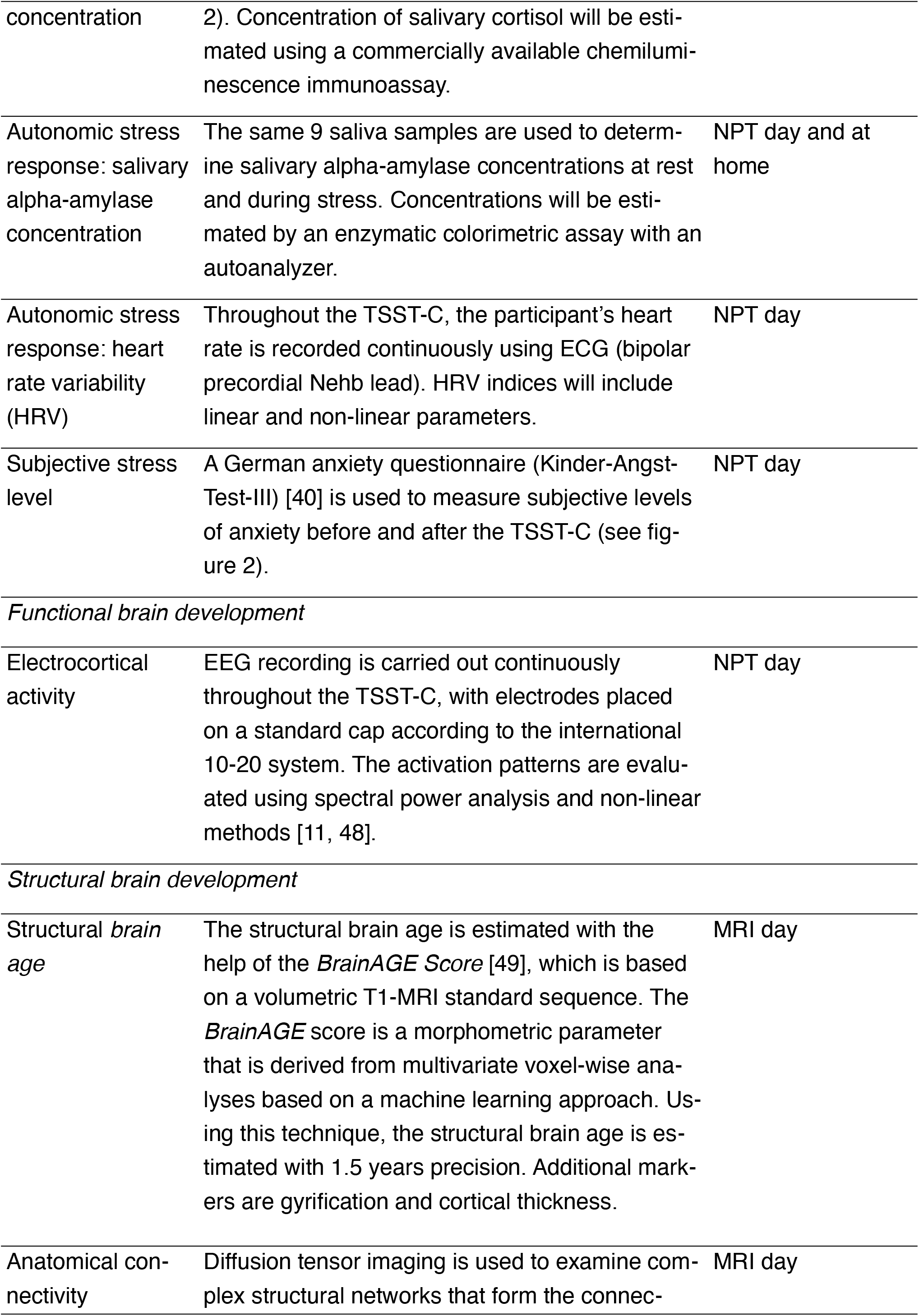
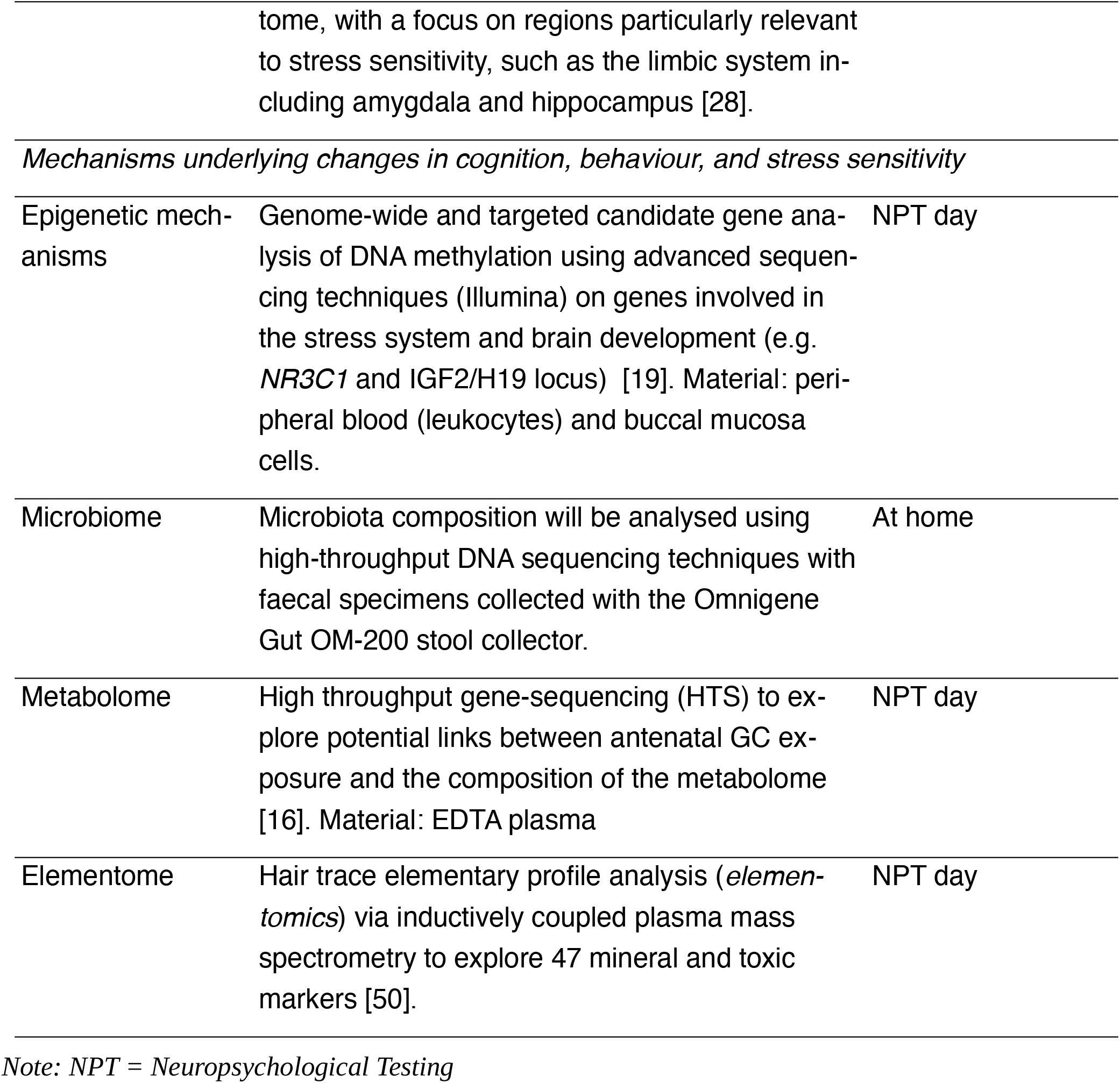
Primary and secondary outcome measures.

| Variables | Instruments/Scale | Collection |
| --- | --- | --- |
| **Primary outcome measure** |  |  |
| Intelligence quotient (IQ) | General cognitive ability is examined using the German version of the Reynolds Intellectual Assessment Scales and Screening (RIAS) [37]. It examines facets of verbal and nonverbal intelligence and memory capacity. | NPT day |
| **Secondary outcome measures** |  |  |
| *Cognition and behaviour of the child* |  |  |
| Selective attention, vigilance, impulsivity | A continuous performance test (CPT) [42] is used to assess attentional performance in terms of selective attention, vigilance, and impulsivity. | NPT day |
| Motor development | To examine motor development, the Movement Assessment Battery for Children – Second Edition (M-ABC-2) [43] is used, which assesses the developmental adequacy of manual dexterity, ball skills, and balance. | NPT day |
| Emotional excitability | To assess emotional excitability, the corresponding scale of a German self-report personality assessment for children [44] is used. | NPT day |
| Attention deficit disorder symptoms (attention, hyperactivity, and impulsivity) | A German parent-reported questionnaire is used to assess attention deficit hyperactivity disorder associated behaviour (*FBB-ADHS* from *DISYPS-III*) [45]. | NPT day |
| Behavioural difficulties I | The German version of the parent-reported Child Behaviour Checklist for children (*CBCL/6-18R*) [46] is used to examine behavioural symptoms on eight scales - (1) anxious/depressed, (2) withdrawn/depressed, (3) somatic complaints, (4) social problems, (5) thought, sleep, and repetitive behaviour problems, (6) attention problems, (7) rule-breaking behaviour, and (8) aggressive behaviour. | NPT day |
| Behavioural difficulties II | A parent-reported screening questionnaire, the German version of the Strengths and Difficulties Questionnaire [47] is used to assess emotional and behavioural problems, hyperactivity and inattention, peer relationship problems, and prosocial behaviour. | NPT day |
| *Stress reactivity during the Trier Social Stress Test for children (TSST-C)* |  |  |
| Neuroendocrine stress response: salivary cortisol | To determine HPAA activity at rest and during stress, a total of 9 saliva samples are collected throughout the TSST-C and at home (see figure | NPT day and at home |
| concentration | 2). Concentration of salivary cortisol will be estimated using a commercially available chemiluminescence immunoassay. |  |
| Autonomic stress response: salivary alpha-amylase concentration | The same 9 saliva samples are used to determine salivary alpha-amylase concentrations at rest and during stress. Concentrations will be estimated by an enzymatic colorimetric assay with an autoanalyzer. | NPT day and at home |
| Autonomic stress response: heart rate variability (HRV) | Throughout the TSST-C, the participant's heart rate is recorded continuously using ECG (bipolar precordial Nehb lead). HRV indices will include linear and non-linear parameters. | NPT day |
| Subjective stress level | A German anxiety questionnaire (Kinder-Angst-Test-III) [40] is used to measure subjective levels of anxiety before and after the TSST-C (see figure 2). | NPT day |
| *Functional brain development* |  |  |
| Electrocortical activity | EEG recording is carried out continuously throughout the TSST-C, with electrodes placed on a standard cap according to the international 10-20 system. The activation patterns are evaluated using spectral power analysis and non-linear methods [11, 48]. | NPT day |
| *Structural brain development* |  |  |
| Structural *brain age* | The structural brain age is estimated with the help of the *BrainAGE Score* [49], which is based on a volumetric T1-MRI standard sequence. The *BrainAGE* score is a morphometric parameter that is derived from multivariate voxel-wise analyses based on a machine learning approach. Using this technique, the structural brain age is estimated with 1.5 years precision. Additional markers are gyrification and cortical thickness. | MRI day |
| Anatomical connectivity | Diffusion tensor imaging is used to examine complex structural networks that form the connec- | MRI day |
|  | tome, with a focus on regions particularly relevant to stress sensitivity, such as the limbic system including amygdala and hippocampus [28]. |  |
| *Mechanisms underlying changes in cognition, behaviour, and stress sensitivity* |  |  |
| Epigenetic mechanisms | Genome-wide and targeted candidate gene analysis of DNA methylation using advanced sequencing techniques (Illumina) on genes involved in the stress system and brain development (e.g. *NR3C1* and IGF2/H19 locus) [19]. Material: peripheral blood (leukocytes) and buccal mucosa cells. | NPT day |
| Microbiome | Microbiota composition will be analysed using high-throughput DNA sequencing techniques with faecal specimens collected with the Omnigene Gut OM-200 stool collector. | At home |
| Metabolome | High throughput gene-sequencing (HTS) to explore potential links between antenatal GC exposure and the composition of the metabolome [16]. Material: EDTA plasma | NPT day |
| Elementome | Hair trace elementary profile analysis (*elementomics*) via inductively coupled plasma mass spectrometry to explore 47 mineral and toxic markers [50]. | NPT day |

*Note: NPT = Neuropsychological Testing*

### Procedure

Based on logistical considerations, eligible participants are invited to two study days (see figure 1). On one of the days, neuropsychological testing (NPT) and the TSST-C take place (NPT day), while an MRI scan is carried out on the other (MRI day). For scheduling reasons, a few days may lie in between visits.

**Figure 1:**
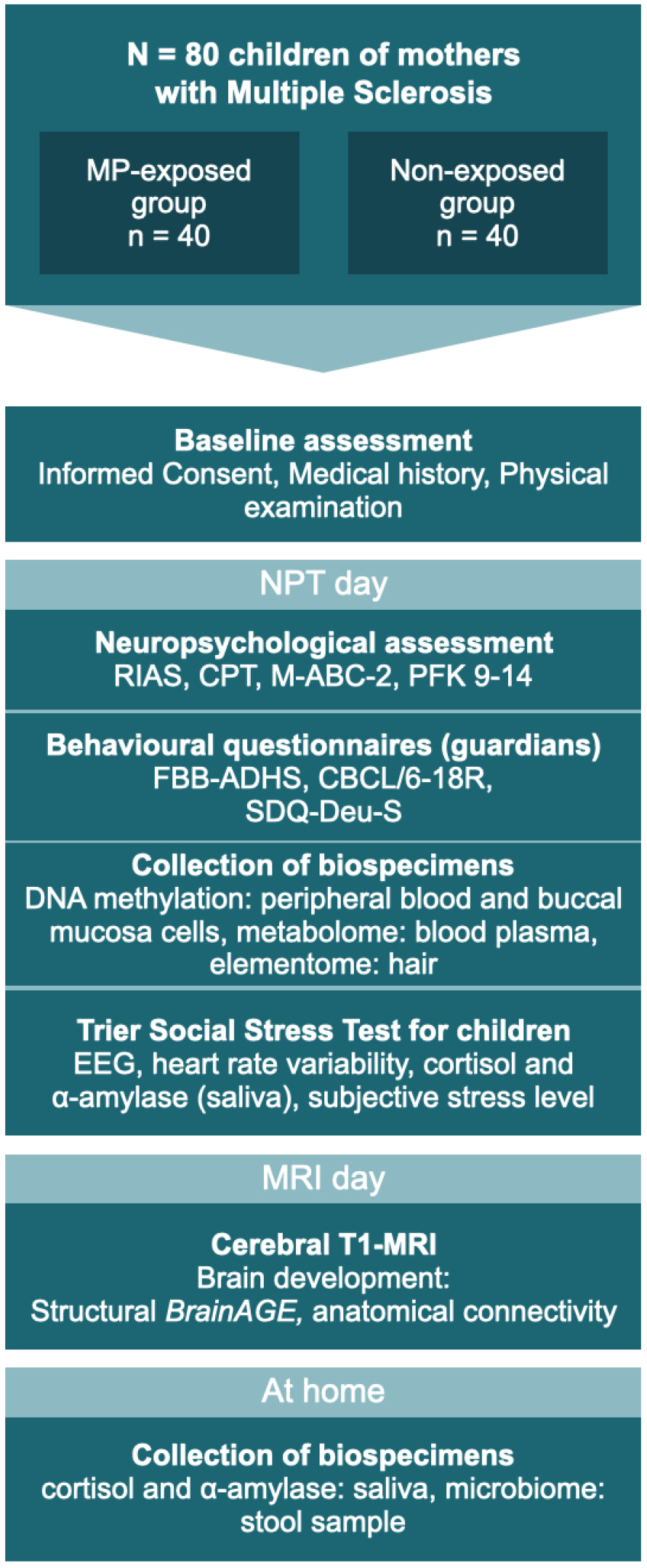
The two study days: testing sequence for all participants. MP = methylprednisolone; NPT = Neuropsychological testing; RIAS = Reynolds Intellectual Assessment Scales and Screening; CPT = Continuous Performance Test; M-ABC-2 = Movement Assessment Battery for Children – Second Edition; PFK 9-14 = German personality questionnaire; FBB-ADHS = German questionnaire for attention deficit hyperactivity disorder; CBCL/6-18 = Child Behaviour Checklist for children aged 6–18 years; SDQ-Deu-S = Strengths and Difficulties Questionnaire. MRI and NPT days may occur in reverse order.

On the NPT day, basic clinical and demographic data are recorded and the accompanying guardian is asked to assess their child’s behaviour using several well-established questionnaires (see tables 1 and 2). The participating child takes part in a set of four neuropsychological assessments of cognition and behaviour (see table 2), performed by a psychologist blinded to the participant’s status of antenatal MP exposition. Before the participant is released for a 60-minute break, hair, saliva, and buccal specimens are collected. The entire procedure takes approximately 120 – 180 minutes.

Following the collection of blood (see table 2), stress sensitivity is examined using the TSST-C, an instrument to generate a psychophysiological stress response in the standardised setting of a simulated examination. See figure 2 for a schematic representation of the procedure. The participant is equipped with EEG and ECG electrodes, a blood pressure monitor, and a pulse oximeter for continuous measurements during the TSST-C. Saliva samples are collected to measure cortisol and alpha-amylase concentrations throughout the test (see figure 2 and table 2 for scheduling of sampling). The TSST-C begins with an ‘active rest’ phase of 15 minutes, during which the participant watches a slow-paced animal documentary, sitting down. In a separate room, a two-person examination committee then present the participant with a story, before leaving the room for 5 minutes, giving the participant time to prepare an ‘exciting and compelling’ ending to story. The participant is then asked to continue telling the story (stress phase I), after which the participant is instructed to verbalise serial subtractions of age-appropriate difficulty (stress phase II). When the participant makes a mistake, the examiner interrupts them with a prompt to start over. The stress phases last a combined 10 minutes, after which the participant receives positive feedback from the committee. In total, the TSST-C takes approximately 130 minutes. After the TSST-C, the participants are handed a kit for the at-home collection of a further 3 saliva samples, to capture the HPAA activity throughout a normal day. The kit also includes material to collect a stool sample for the analysis of the microbiome.

**Figure 2:**
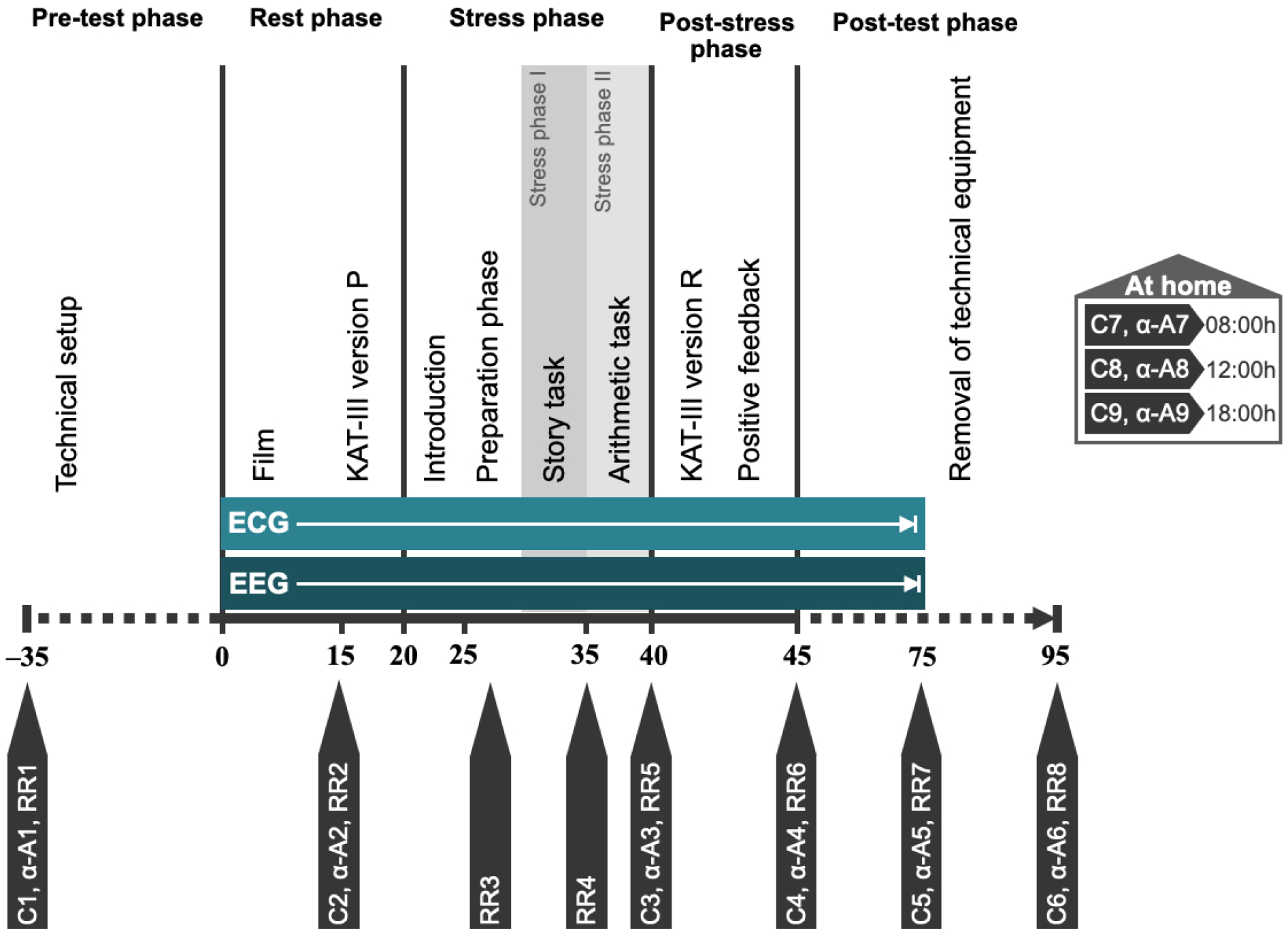
Schematic representation of TSST-C procedure. C1–C9: saliva swabs 1–9 (cortisol); α-A1–α-A9: saliva swabs 1–9 (alpha-Amylase); RR1–RR6: blood pressure measurements 1–6; KAT-III: German anxiety scale for children (Kinder-Angst-Test-III) – versions P (prospective) and R (retrospective).

On the MRI day, a volumetric cranial MRI scan in T1 standard sequence is obtained from the participant. Diffusion tensor imaging (DTI) is used to determine the anatomical connectome. Including participant education, the entire procedure is estimated to take 50 minutes.

### Data storage and management

All data are recorded in a pseudonymised form. In accordance with EU data protection legislation, both study centres maintain a secured patient identification list with full names, addresses, and dates of birth. Any personal information is archived for at least ten years after inclusion of the last participant.

The collection, management and use of human biological specimens in this study is in accordance with national and EU legislation. Any transfer of specimens between the two centres will take place in accordance with the Oviedo Convention (Convention on Human Rights and Biomedicine – CETS 164) and Recommendation of the Committee of Ministers to Member States on research on biological materials of human origin (adopted by the Committee of Ministers on 15 March 2006 at the 958th Meeting of Ministerial Deputies). Due to the vulnerable nature of a cohort of children, we further implement directive 95/46/EC and working group 8/2010 opinion, article 29 and take the high importance of data protection in vulnerable populations into careful consideration.

Part of the initial data collection uses hard copy files, which are stored in a secure location and later digitised using an electronic case report form. All digitised data are stored on a safe server managed by the Centre for Clinical Studies at the University hospital in Jena.

### Data analysis plan

For the analysis of the primary outcome, exposed and non-exposed groups will be compared in terms of their performance on the IQ test using a linear regression model taking into account potential confounders, including antenatal exposure to MP, point in time of exposure, maternal age at birth, birth weight, and socioeconomic background. Exploratory analysis of secondary outcomes will further be performed with a linear regression model. Those outcomes measured repeatedly throughout the course of the TSST-C will be evaluated as time series data with a linear mixed model, with time as a further independent factor. For this analysis, two models will be fitted to explore (1) overall mean activity throughout the TSST-C, as well as (2) the change from baseline, in which the stress response will be adjusted to the baseline value.

### Methodological challenges

Recruitment of eligible participants presents the main challenge for this study. However, based on estimations of the prevalence of MS in Germany as well as pregnancy and relapse rates, our sample size represents only approximately .8% of the total target population in Germany. The close cooperation with the *German MS and Pregnancy Registry*, founded by members of the study group, gives us access to a database of more than 3,000 pregnancies of women with MS. Several MS clinics and surgeries across Germany, as well as the German Society for MS (DMSG) support recruitment efforts by sharing flyers and online adverts with their patients or members, and searching their databases for potential participants. The ongoing COVID-19 pandemic presents another challenge which slowed down recruitment and testing, particularly in the first quarter of 2021. In accordance with the rules and recommendations of the German government, we have implemented a strict hygiene plan for participants and researchers to continue with recruitment.

The TSST-C is vulnerable to subjective factors in the examination committee and environmental factors. The procedure is guided by detailed protocols and pilot runs, to ensure standardisation within and between study centres. The same investigator performs neuropsychological testing in both study centres, granting further consistency of conditions.

To account for the vulnerability of the study population and potential anxiety around measures such as MRI or blood sampling, we devised detailed, child-friendly participant education material. The children’s guardians are usually present and the child is offered a topical local anaesthetic in preparation for the drawing of blood.

In this study, antenatal or postnatal stressors may act as confounds. Maternal psychosocial stress during pregnancy has been shown to elicit increased maternal and foetal cortisol levels [51, 52]. It is further associated with an increase in the offspring stress response, disturbed motor and structural brain development and an increased risk of cognitive, behavioural, and emotional problems in later life, such as autism spectrum disorders, ADHD, depression and schizophrenia (for reviews see [52, 53]). MS relapses are not only treated with GCs but also are a major psychological stressor. However, as relapses during pregnancy rarely go untreated, there is no realistic way of mitigating this confounder using an untreated MS control group. To take this into account, participants and their mothers fill out questionnaires regarding subjective and objective stress during pregnancy and the past 12 months of the participant’s life (see table 1).

## ETHICS AND DISSEMINATION

This study is conducted with the approval of the ethics committees of Friedrich-Schiller University Jena (reference: 2020-1668-3-BO) and the medical faculty of Ruhr University Bochum (reference: 21-7192 BR). The results are going to be published in relevant peer-reviewed journals, presented in accordance with the STROBE 2007 Statement [54].

## Data Availability

All data produced in this study will be made available in an appropriate online data repository.

## Author contributions

VK: writing of the manuscript, study implementation, data collection; MS: study conception and design, critical manuscript revision; ST and KS: critical manuscript revision, study implementation; FR: study conception, design, study implementation, critical manuscript revision; MD: study conception, design, critical manuscript revision, study implementation, data collection

## Acknowledgements

We would like to thank Dr. phil. Carolin Ligges for her expertise and council regarding psychological testing and research with children.

## References

1. Browne, P., et al., Atlas of Multiple Sclerosis 2013: A growing global problem with widespread inequity. Neurology, 2014. 83(11): p. 1022–1024.

2. Kingwell, E., et al., Incidence and prevalence of multiple sclerosis in Europe: a systematic review. BMC neurology, 2013. 13(1): p. 1–13.

3. Thöne, J., et al., Treatment of multiple sclerosis during pregnancy – safety considerations. Expert Opinion on Drug Safety, 2017. 16(5): p. 523–534.

4. Confavreux, C., et al., Rate of pregnancy-related relapse in multiple sclerosis. New England Journal of Medicine, 1998. 339(5): p. 285–291.

5. Dobson, R., et al., UK consensus on pregnancy in multiple sclerosis:’Association of British Neurologists’ guidelines. Practical neurology, 2019. 19(2): p. 106–114.

6. Lockshin, M.D. and L.R. Sammaritano, Corticosteroids during pregnancy. Scandinavian Journal of Rheumatology, 1998. 27(Sup107): p. 136–138.

7. Alexander, N., et al., Impact of antenatal synthetic glucocorticoid exposure on endocrine stress reactivity in term-born children. The Journal of Clinical Endocrinology & Metabolism, 2012. 97(10): p. 3538–3544.

8. Ilg, L., et al., Persistent Effects of Antenatal Synthetic Glucocorticoids on Endocrine Stress Reactivity From Childhood to Adolescence. The Journal of Clinical Endocrinology & Metabolism, 2019. 104(3): p. 827–834.

9. Liu, D., et al., A practical guide to the monitoring and management of the complications of systemic corticosteroid therapy. Allergy, Asthma & Clinical Immunology, 2013. 9(1): p. 30.

10. Rakers, F., et al., Impact of antenatal glucocorticoid exposure on activity of the stress system, cognition and behavior in 8–9-year-old children: a clinical cohort study. In preparation.

11. Schwab, M., et al., Effects of Prenatal Betamathasone (BM) Exposure on Cortical Activity at the Age of Eight Years-A Pilot Study. Reproductive Sciences, 2010. 17(3): p. 292A–293A.

12. Berger, R., et al., Prävention und Therapie der Frühgeburt. Leitlinie der DGGG, OEGGG und SGGG (S2k-Niveau, AWMF-Registernummer 015/025, Februar 2019)–Teil 1 mit Empfehlungen zur Epidemiologie, Ätiologie, Prädiktion, primären und sekundären Prävention der Frühgeburt. Zeitschrift für Geburtshilfe und Neonatologie, 2019. 223(05): p. 304–316.

13. Berger, R., et al., Prävention und Therapie der Frühgeburt. Leitlinie der DGGG, OEGGG und SGGG (S2k-Niveau, AWMF-Registernummer 015/025, Februar 2019)–Teil 2 mit Empfehlungen zur tertiären Prävention der Frühgeburt und zum Management des frühen vorzeitigen Blasensprungs. Zeitschrift für Geburtshilfe und Neonatologie, 2019. 223(06): p. 373–394.

14. Barker, D.J., The fetal and infant origins of adult disease. BMJ: British Medical Journal, 1990. 301(6761): p. 1111.

15. Gillman, M.W., Developmental origins of health and disease. The New England journal of medicine, 2005. 353(17): p. 1848.

16. Rakers, F., et al., Transfer of maternal psychosocial stress to the fetus. Neuroscience & Biobehavioral Reviews, 2020. 117: p. 185–197.

17. Benediktsson, R., et al., Placental 11β-hydroxysteroid dehydrogenase: a key regulator of fetal glucocorticoid exposure. Clinical endocrinology, 1997. 46(2): p. 161–166.

18. Anderson, G.G., Y. Rotchell, and D.G. Kaiser, Placental transfer of methylprednisolone following maternal intravenous administration. American journal of obstetrics and gynecology, 1981. 140(6): p. 699–701.

19. Cao-Lei, L., et al., Prenatal stress and epigenetics. Neuroscience & Biobehavioral Reviews, 2020. 117: p. 198–210.

20. Harris, A. and J. Seckl, Glucocorticoids, prenatal stress and the programming of disease. Hormones and Behavior, 2011. 59(3): p. 279–289.

21. Moisiadis, V.G. and S.G. Matthews, Glucocorticoids and fetal programming part 1: Outcomes. Nature Reviews Endocrinology, 2014. 10(7): p. 391.

22. Liggins, G., The role of cortisol in preparing the fetus for birth. Reproduction, Fertility and Development, 1994. 6(2): p. 141–150.

23. Garbrecht, M.R., et al., Glucocorticoid metabolism in the human fetal lung: implications for lung development and the pulmonary surfactant system. Biology of the Neonate, 2006. 89(2): p. 109–119.

24. Bloom, S.L., et al., Antenatal dexamethasone and decreased birth weight. Obstetrics & Gynecology, 2001. 97(4): p. 485–490.

25. Uno, H., et al., Neurotoxicity of glucocorticoids in the primate brain. Hormones and Behavior, 1994. 28(4): p. 336–348.

26. Carson, R., et al., Effects of antenatal glucocorticoids on the developing brain. Steroids, 2016. 114: p. 25–32.

27. Antonow-Schlorke, I., et al., Adverse effects of antenatal glucocorticoids on cerebral myelination in sheep. Obstetrics & Gynecology, 2009. 113(1): p. 142–151.

28. Scheinost, D., et al., Does prenatal stress alter the developing connectome? Pediatric research, 2017. 81(1): p. 214–226.

29. Huang, E.Y., et al., Using corticosteroids to reshape the gut microbiome: implications for inflammatory bowel diseases. Inflammatory bowel diseases, 2015. 21(5): p. 963–972.

30. Zhao, H., X. Jiang, and W. Chu, Shifts in the gut microbiota of mice in response to dexamethasone administration. International Microbiology, 2020. 23(4): p. 565–573.

31. Jašarević, E., et al., The maternal vaginal microbiome partially mediates the effects of prenatal stress on offspring gut and hypothalamus. Nature neuroscience, 2018. 21(8): p. 1061–1071.

32. Vuong, H.E., et al., The maternal microbiome modulates fetal neurodevelopment in mice. Nature, 2020. 586(7828): p. 281–286.

33. Glover, V., et al., Prenatal maternal stress, fetal programming, and mechanisms underlying later psychopathology—a global perspective. Development and psychopathology, 2018. 30(3): p. 843–854.

34. Vuong, H.E., et al., The microbiome and host behavior. Annual review of neuroscience, 2017. 40: p. 21–49.

35. Baker, J., et al., Role of insulin-like growth factors in embryonic and postnatal growth. Cell, 1993. 75(1): p. 73–82.

36. Austin, P.C. and E.W. Steyerberg, The number of subjects per variable required in linear regression analyses. Journal of Clinical Epidemiology, 2015. 68(6): p. 627–636.

37. Hagmann-Von, A. and A. Grob, Reynolds Intellectual Assessment Scales and Screening. Deutschsprachige Adaptation der Reynolds Intellectual Assessment Scales (RIAS) & des Reynolds Intellectual Screening Test (RIST) von Cecil R. Reynolds und Randy W. Kamphaus. 2014, Bern: Hans Huber, Hogrefe AG.

38. Kirschbaum, C., K.-M. Pirke, and D.H. Hellhammer, The ‘Trier Social Stress Test’–a tool for investigating psychobiological stress responses in a laboratory setting. Neuropsychobiology, 1993. 28(1-2): p. 76–81.

39. Nater, U.M. and N. Rohleder, Salivary alpha-amylase as a non-invasive biomarker for the sympathetic nervous system: current state of research. Psychoneuroendocrinology, 2009. 34(4): p. 486–496.

40. Tewes, A. and A. Naumann, Kinder-Angst-Test III. 1 ed. 2016, Göttingen: Hogrefe.

41. Steinhausen, H.-C. and C. Winkler Metzke, Die Zürcher Lebensereignis-Liste (ZLEL): Ergebnisse einer Schweizer epidemiologischen Untersuchung. Kindheit und Entwicklung, 2001. 10(1): p. 47–55.

42. Knye, M., et al., Continuous Performance Test: CPT. 1 ed. 2004, Göttingen: Hogrefe.

43. Henderson, S.E., D.A. Sudgen, and A.L. Barnett, Movement Assessment Battery for Children -2 (M-ABC-2; German edition). 4 ed, ed. F. Petermann. 2015: Pearson Assessment.

44. Seitz, W. and A. Rausche, Persönlichkeitsfragebogen für Kinder zwischen 9 und 14 Jahren: PFK 9-14. 5 ed. 2004, Göttingen: Hogrefe.

45. Döpfner, M. and A. Görtz-Dorten, Diagnostik-System für psychische Störungen nach ICD-10 und DSM-5 für Kinder-und Jugendliche (DISYPS-III). 1 ed. 2017, Bern: Hogrefe.

46. Döpfner, M., et al., Deutsche Schulalter-Formen der Child Behavior Checklist von Thomas M. Achenbach: Elternfragebogen über das Verhalten von Kindern und Jugendlichen (CBCL/6-18R). 1 ed. 2014, Göttingen: Hogrefe.

47. Lohbeck, A., et al., Die Deutsche selbstbeurteilungsversion des strengths and difficulties questionnaire (SDQ-Deu-S). Diagnostica, 2015.

48. Schmidt, K., et al., Nonlinear modeling of different fetal and neonatal behaviorial states. Pathophysiology, 1998. 1001(5): p. 257.

49. Franke, K. and C. Gaser, Ten years of BrainAGE as a neuroimaging biomarker of brain aging: what insights have we gained? Frontiers in neurology, 2019. 10: p. 789.

50. Ambeskovic, M., et al., Hair trace elementary profiles in aging rodents and primates: links to altered cell homeodynamics and disease. Biogerontology, 2013. 14(5): p. 557–567.

51. Rakers, F., et al., Role of catecholamines in maternal-fetal stress transfer in sheep. American journal of obstetrics and gynecology, 2015. 213(5): p. 684. e1–684. e9.

52. Van den Bergh, B.R., R. Dahnke, and M. Mennes, Prenatal stress and the developing brain: Risks for neurodevelopmental disorders. Development and psychopathology, 2018. 30(3): p. 743–762.

53. Van den Bergh, B.R., et al., Prenatal developmental origins of behavior and mental health: The influence of maternal stress in pregnancy. Neuroscience & Biobehavioral Reviews, 2020. 117: p. 26–64.

54. Von Elm, E., et al., The Strengthening the Reporting of Observational Studies in Epidemiology (STROBE) statement: guidelines for reporting observational studies. Bulletin of the World Health Organization, 2007. 85: p. 867–872.

